# Evaluating the relationship between glycemic control and bone fragility within the UK biobank: Observational and one-sample Mendelian randomization analyses

**DOI:** 10.1101/2023.12.25.23300434

**Authors:** Samuel Ghatan, Fjorda Koromani, Katerina Trajanoska, Evert F.S. van Velsen, Maryam Kavousi, M Carola. Zillikens, Carolina Medina-Gomez, Ling Oei, Fernando Rivadeneira

## Abstract

**Aims/hypothesis:** This study aimed to: (1) examine the relationship between glycemic control, bone mineral density estimated from heel ultrasound (eBMD) and fracture risk in individuals with type 1 and type 2 diabetes and (2) perform a one-sample Mendelian randomization study to explore potential linear and non-linear associations between glycemic control, eBMD, and fractures.

**Methods:** This study comprised 452,131 individuals from the UK Biobank with glycated hemoglobin A1C (HbA1c) and eBMD levels. At baseline, 4,078 participants were diagnosed with type 1 diabetes and 23,682 with type 2 diabetes. HbA1c was used to classify patients into “adequately-” (ACD; n=17,078; HbA1c < 7.0%/53mmol/mol) and “inadequately-” (ICD; n=10,682; HbA1c ≥ 7.0%/53mmol/mol) controlled diabetes. The association between glycemic control (continuous and categorical) and eBMD was tested using linear regression, while fracture risk was estimated in Cox regression models, both controlling for covariates. Mendelian randomization (MR) was used to evaluate linear and non-linear causal relationships between HbA1c levels, fracture risk, and eBMD.

**Results:** In individuals with type 1 diabetes, a 1% unit (11mmol/mol) increase in HbA1c levels was associated with a 12% increase in fracture risk (HR: 1.12, 95% CI [1.05-1.19]). Individuals with type 1 diabetes had lower eBMD in both the ICD (beta = −0.08, 95% CI [−0.11, −0.04]) and ACD (beta = −0.05, 95% CI [-0.11,0.01]) groups, as compared to subjects without diabetes. Fracture risk was highest in individuals with type 1 diabetes and ICD (HR 2.84, 95%CI [2.53, 3.19]), followed by those with ACD (HR 2.26, 95%CI [1.91, 2.69]). Individuals with type 2 diabetes had higher eBMD in both ICD (beta=0.12SD, 95%CI [0.10, 0.14]) and ACD (beta=0.07SD, 95%CI [0.05, 0.08]) groups. Significant evidence for a non-linear association between HbA1c and fracture risk was observed (F-test ANOVA p-value = 0.002) in individuals with type 2 diabetes, with risk being increased at both low and high levels of HbA1c. Fracture risk between the type 2 diabetes ACD and ICD groups was not significantly different (HR: 0.97, 95%CI [0.91-1.16]), despite increased BMD. In MR analyses genetically predicted higher HbA1c levels were not significantly associated with fracture risk (Causal Risk Ratio: 1.04, 95%CI [0.95-1.14]). However, disease stratified analyses were underpowered. We did observe evidence of a non-linear causal association with eBMD (quadratic test P-value = 0.0002), indicating U-shaped relationship between HbA1c and eBMD.

**Conclusion/interpretation:** We obtained evidence that lower HbA1c levels will reduce fracture risk in patients with type 1 diabetes. In individuals with type 2 diabetes, lowering HbA1c levels can mitigate the risk of fractures up to a threshold, beyond which the risk may begin to rise once more. MR analyses demonstrated a causal relationship between genetically predicted HbA1c levels and eBMD, but not fracture risk.

## Introduction

Diabetes mellitus is a common metabolic disorder characterized by elevated blood glucose levels, affecting millions of individuals worldwide. While the detrimental effects of poor glycemic control in diabetes on various organ systems have been extensively investigated [1], its impact on skeletal health remains an area of ongoing research. Previous studies have established fracture risk as a complication of diabetes [2-4]. Fractures pose significant challenges, leading to considerable morbidity, mortality, and healthcare costs [5, 6]. However, understanding the complex relation between glycemic control, measured by glycated hemoglobin (HbA1c) levels, and fracture risk in individuals with both type 1 diabetes and type 2 diabetes is crucial for implementing preventive measures and improving patient outcomes [7]. Additionally, investigating the relationship between glycemic control and bone mineral density (BMD), a key determinant of fracture susceptibility, can offer insights into potential therapeutic strategies for mitigating the skeletal complications associated with diabetes. Glycemic control has been shown to be a key determinant of other complications of diabetes including microvascular disease, myocardial infarction and all-cause mortality [1]. To address these knowledge gaps comprehensively, we conducted a large-scale observational study utilizing data from the UK Biobank, a well-characterized population-based resource [8]. Our study aimed to examine the relationship between glycemic control, BMD and fracture risk in individuals with type 1 diabetes and type 2 diabetes; and utilize linear and non-linear Mendelian randomization to explore potential causal associations.

### Research Design and Methods

#### Study population

The UK biobank is a prospective cohort study that recruited up to 502,410 participants across 22 centers in the UK. Participants were aged between 40-59 years at baseline [8]. HbA1c levels were measured in 482,253 individuals across two center visits. The initial assessment visit took place between 2006-2010 and encompassed 95% of the measurements taken. The second assessment visit took place between 2012-2013. The date of HbA1c measurement was established as baseline. Individuals with prevalent fractures at baseline were excluded (fracture definition provided below). The median follow-up time was 11.72 years during which 20,414 incident fractures occurred. The follow-up time end date was determined as the date of the last recorded fracture. Individuals who did not fracture or died before the end date were censored. The date of an individual’s death was obtained via death registry (data field 40000). A subset of 452,131 individuals also had eBMD measurements available, obtained over three center visits. The initial visit took place between 2006-2010 encompassing 90% of the measurements taken. Subsequent visits took place between 2012-2014 and 2014-2016.

The Mendelian randomization study was performed on a subset of 379,600 individuals of European ancestry who had genotype information available [9]. The UK biobank was conducted in full accordance with the ethical principles outlined in the Declaration of Helsinki, as revised in 2013, including adherence to protocols approved by their respective institutional ethics review committees and all participants provided written informed consent. Diabetes definition and glycemic measures Individuals with type 2 diabetes were identified either via self-reporting at baseline (data field 130709), obtaining a HbA1c level > 6.5% (48 mmol/mol) at baseline, or via a recorded International Classification of Diseases, Tenth Revision (ICD-10) code E11 (non-insulin-dependent diabetes mellitus) (data field 130708). Similarly, individuals with type 1 diabetes were defined using self-reported data, as well as, the ICD-10 code E10 (Insulin-dependent diabetes mellitus). Plasma HbA1c levels were measured using variant II turbo Hemoglobin Testing System; from Bio-Rad. Individuals with diabetes and HbA1c levels ≥ 7% (53mmol/mol) were considered to have inadequate glycemic control, conversely those with <7% were considered as having adequate control, according to American Diabetes Association (ADA) guidelines [10]. The duration of diabetes was determined based on either the self-reported age of diabetes diagnosis (data field 2976) or the earliest date of the corresponding ICD-10 code. In cases where self-reported ages were unavailable or entered incorrectly, they were replaced with the first date of the ICD-10 code. Pre-diabetes was defined using ADA guidelines as HbA1c levels of 5.7-6.4% (39-46mmol/mol) [11].

#### Fractures and BMD measurements

Estimated BMD (eBMD) measurements were calculated from measured quantitative ultrasound speed of sound (SOS) and broadband ultrasound attenuation (BUA) of the heel. Measurements were collected over three timepoints. The data collection and quality control of eBMD measurements were conducted in accordance with the procedure outlined by the GEFOS consortium [12]. Fractures were defined according to ICD-10 codes. A full list of the codes used can be located in supplementary table 1. Fractures of the skull, face, hands and feet, atypical femoral fractures, pathological fractures due to malignancy, periprosthetic, and healed fracture codes were excluded. Individuals who retracted their informed consent, as of May 4^th^, 2023, were removed.

#### Complications

Cardiovascular disease was defined using the ICD-10 codes I20, angina pectoris; I21, acute myocardial infarction; 122 subsequent ST elevation (STEMI) and non-ST elevation (NSTEMI) myocardial infarction; I23, certain current complications following ST elevation (STEMI) and non-ST elevation (NSTEMI) myocardial infarction; I24, other acute ischemic heart diseases; or I25, chronic ischemic heart disease. Chronic kidney disease was defined by ICD-10 code N18; Chronic kidney disease. Diabetic retinopathy was defined by the ICD-10 codes: E08.3 -Diabetes mellitus due to underlying condition with ophthalmic complications, E09.3 - Drug or chemical induced diabetes mellitus with ophthalmic complications, E10.3 - Type 1 diabetes mellitus with ophthalmic complications, E11.3 - Type 2 diabetes mellitus with ophthalmic complications, E13.3 - Other specified diabetes mellitus with ophthalmic complications.

### Statistical analyses

#### Observational associations

The reporting of epidemiological results was undertaken following the STROBE and STROBE-MR guidelines. The effect of HbA1C on BMD and fracture risk was estimated from linear regression and Cox proportional hazards models, respectively. Estimates were also obtained for the association between glycemic groups (i.e., ACD, ICD, pre-diabetes) and outcomes in comparison to diabetes-free individuals. Additionally, to compare the fracture risk between glycemic groups hazard ratios (HR) were obtained when setting ACD as the reference group. The continuous effect of HbA1c levels on eBMD and fractures was assessed in type 1 diabetes, type 2 diabetes and diabetes-free sub-group analyses. Non-linear effects were also evaluated using linear tail-restricted cubic spline models with 3 knots using the ‘rms’ package in R. The number of knots was determined by visual inspection of plots and using Akaike information criterion, to compare models with different degrees of freedom. The significance of the non-linear spline terms was evaluated using an F-test ANOVA test. All models were adjusted for: age, age^2, sex, height, weight, smoking status (data field 20116), alcohol intake (data field 1558), creatinine (23478), c-reactive protein (data field 30710), menopause (data field 2724), genetic ethnicity (data field 22006), self-reported ethnicity (data field 21000), index of Multiple Deprivation (data field 26410), and duration of moderate activity (data field 894). In Cox models the proportionality of hazards assumption was evaluated using Schoenfeld residual plots. Variables with missing information (less than 20%) were imputed using the mice package in R [13].

#### Sensitivity analyses

For sensitivity analyses, a second higher HbA1c cut-off was defined at 9% (75mmol/mol) with those above it being classified as having adequate control and vice versa. This cut-off was motivated by papers showing increased fracture risk at higher HbA1c cut-offs [14, 15]. Additionally, to investigate the association between low HbA1c and fracture risk, we further split those individuals with type 2 diabetes and ACD by a HbA1c level of below 5.4% (36mmol/mol) (the mean of the sample) and labelled these individuals as hypoglycemic. The risk of fractures in individuals with pre-diabetes was also evaluated. The effect of diabetes duration of fracture risk was also assessed in individuals with type 2 diabetes and available duration data (n=19,263) by including diabetes duration as a covariate in the Cox model. Additionally, disease duration was categorized into 3 groups: less than 5 years, more than 5 years less but than 10 years, and 10 years and greater. Effect estimates were obtained using Cox regression models with the group of less than 5 years as the reference category. Lastly, to assess the potential effect diabetic complications and comorbidities on fracture risk and HbA1c levels we adjusted our statistical models for hemoglobin concentration (data field 30020), as a measure of anemia, chronic kidney disease, cardiovascular disease, and diabetic retinopathy.

#### Selection of genetic instrumental variables

We selected 88 conditionally independent genetic variants as instrumental variables for HbA1c levels (supplementary table 2) based on their association with HbA1c (p<5×10^-8^) in a genome-wide association study of 191,362 individuals of European ancestry [16]. Palindromic SNPs were removed and replaced by linkage disequilibrium proxies (r^2^ > 0.80). Standardized genetic risk scores (GRS) were constructed using the 88 genetic variants for individuals with available genetic data. To test the instrument strength, linear regression models, adjusted for age at baseline, sex, and 10 genetic principal components of ancestry, were constructed. The GRS explained 3% of the variance of HbA1c within the UK biobank population and displayed an F statistic of 1068.

#### Linear Mendelian randomization

For a genetic variant to act as a valid instrumental variable in a MR analysis it had to satisfy the following assumptions: 1) relevance: reliably associated with each of the exposures included in the model; 2) exclusion restriction: the variant needs to be associated with the outcome only through the exposure of interest; and 3) independence: the variant should be conditionally independent of the outcome given the exposure and confounding factors [17]. To obtain linear causal inferences, we applied a two-stage least squares regression (2SLS) for continuous outcomes and structural mean models (SMM) for binary outcomes [18, 19]. Both regression stages were adjusted by age, sex, and the first 10 principal components of genetic ancestry. A sensitivity analysis was performed by removing related individuals (kingship coefficient < 0.0884) and recalculating regression estimates [20]. MR power calculations were derived using the mRnd web tool [21].

#### Non-linear Mendelian randomization

To assess non-linear causal effects we sought to stratify individuals and calculate localized average causal effects (LACE) using the ratio method (instrumental variable outcome association divided by the instrumental variable exposure association) [22]. To stratify individuals we adopted the doubly-ranked method [22], which overcomes an important limitation of the residual method, namely that it assumes that the effect of the genetic instrument on the exposure is constant and linear. Briefly, the doubly-ranked method is a non-parametric approach for exposure stratification, such that, the stratification is not a function of the instrumental variables and each strata has a different average level of the exposure. This involves the initial ranking of individuals based on their level of the instrumental variables, which are then divided into pre-strata. After this, the individuals within each pre-stratum are ranked based on their level of exposure and further divided into strata. Additional test statistics for non-linearity were calculated using the ‘SUMnlmr’ R package [23]. Next, we obtained causal inferences on the shape of the exposure-outcome relationship using fractional polynomials [24]. These polynomials represent the stratification of individuals into either 10 strata or 100 strata. Non-linearity tests included: the fractional polynomial degree test, the fractional polynomial non-linearity test, the quadratic test and Cochran’s Q-test. The fractional polynomial degree test was used to indicate the polynomial degree preference, with a low p-value indicating preference of a degree 2 polynomial. The fractional polynomial non-linearity tests the best-fitting fractional polynomial of a degree 1 against the linear model, with a low p-value indicating a preference of fractional polynomials of degree 1. The quadratic test meta regresses LACE estimates against the mean value of the exposure in each stratum, with a lower p-value indicating non-linearity. Lastly, the Cochran’s Q-test, tests also whether the LACE estimates differ more than expected by chance, with a lower p-value indicating non-linearity [23, 24].

## Results

### Population characteristics

This study comprised 452,131 individuals from the UK biobank cohort study. At baseline, 4,078 (0.9%) individuals had type 1 diabetes and 23,682 (5.2%) had type 2 diabetes. A third group was comprised of individuals without diabetes (n = 424,371) with a median glycated hemoglobin levels (HbA1c) level of 5.4%. A median of HbA1c level of 7.4% was observed in individuals with type 1 diabetes and of 6.6% in those with type 2 diabetes. Individuals with each type of diabetes were stratified into two groups: HbA1c ≥ 7.0%/53mmol/mol defined as Inadequately Controlled Diabetes (ICD; n=10,682); representing 62.9% of those with type 1 diabetes (n = 2,565) and 34.3% of those with type 2 diabetes (n = 8,117); and those with HbA1c <7.0%/53mmol/mol, defined as Adequately Controlled Diabetes (ACD; n =17,078). The median follow-up time was 11.7 years period during which 20,414 incident fractures occurred. The mean age of the participants was 58 years old (SD = 8) with similar proportion of men and women (Table 1). In individuals with type 2 diabetes the mean disease duration was 5.5 years (SD = 5.2) before inclusion at baseline. Estimated bone mineral density (eBMD), derived via heel ultrasound, was measured in all individuals.

**Table 1.**
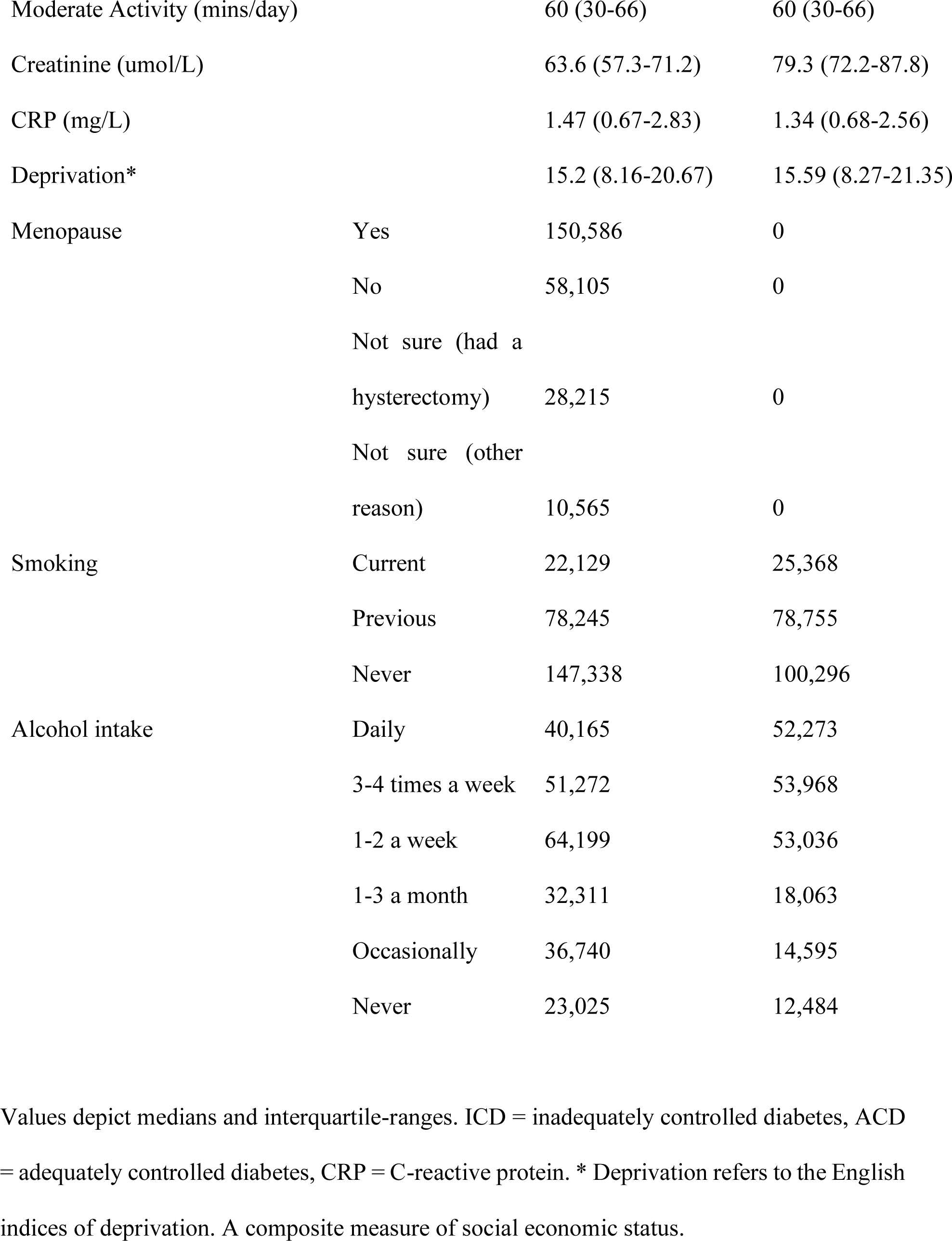
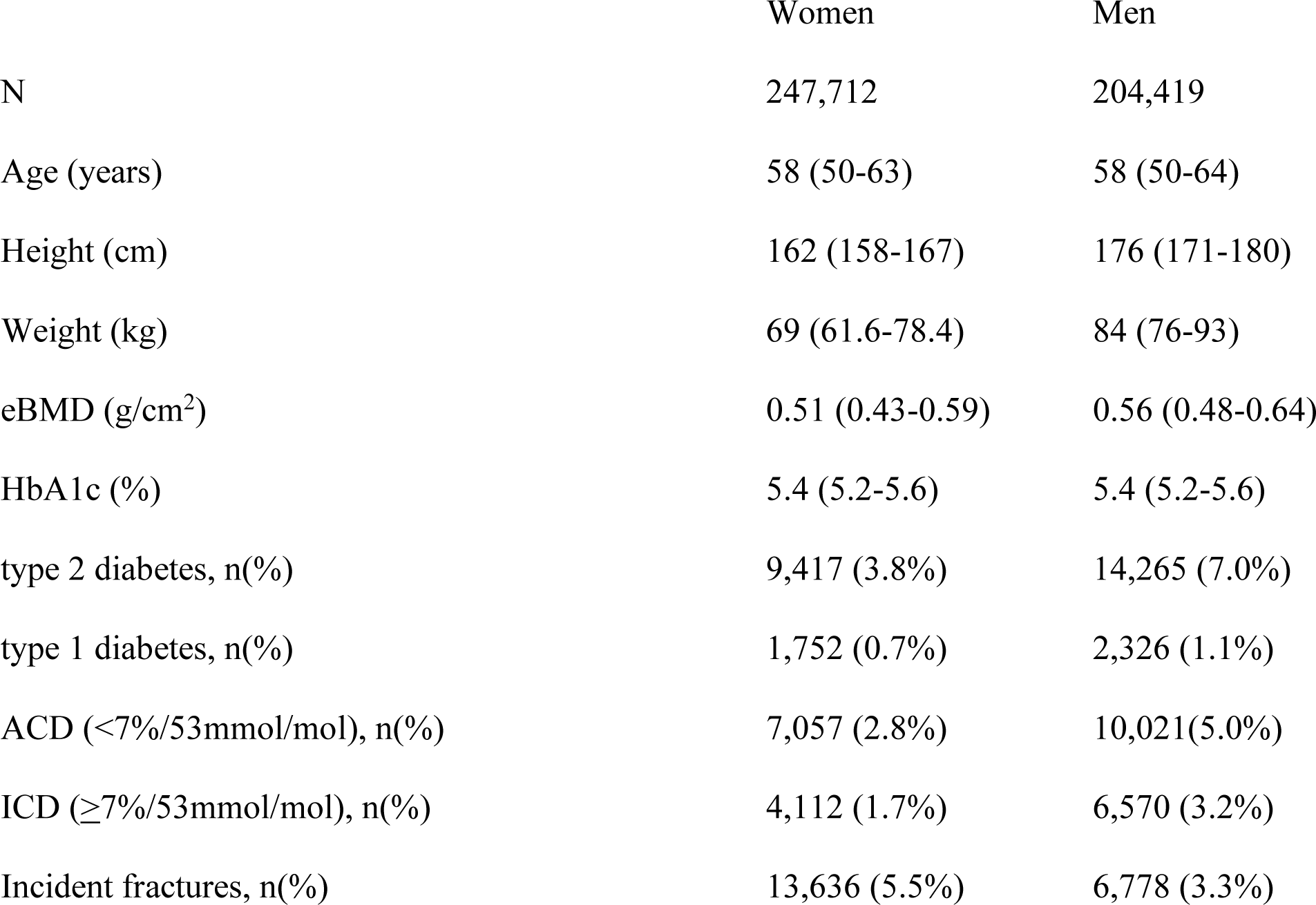
Participant baseline characteristics.

### Glycemic control and estimated Bone Mineral Density

In individuals with type 1 diabetes, HbA1c levels were not significantly associated with eBMD (beta=0.01SD, 95%CI [-0.03, 0.02]) (supplementary table 5), although there was weak evidence suggesting a non-linear U-shaped relationship (F-test ANOVA p-value = 0.08) (Figure 1a). In type 1 diabetes, the ACD (beta=-0.04 SD, 95%CI [-0.08, 0.01]) and ICD (beta=- 0.07 SD, 95%CI [−0.10, −0.02]) groups displayed lower eBMD with respect to HbA1c levels, in comparison to diabetes-free individuals (Figure 2a). In individuals with type 2 diabetes, a 1% (11mmol/mol) increase in HbA1c levels was associated with a 0.01 SD eBMD increase (95%CI [0.00-0.03]) (supplementary table 5). No evidence of a non-linear association between HbA1c and eBMD was observed for type 2 diabetes in sub-group analyses (F-test ANOVA p-value = 0.72) (Figure 1b). In the type 2 diabetes analysis, stratification by glycemic control yielded concordant results with a positive association between HbA1c and eBMD in the ACD (beta=0.07SD, 95%CI [0.05, 0.08]) and ICD (beta=0.12SD, 95%CI [0.10, 0.14]) groups.

**Figure 1.**
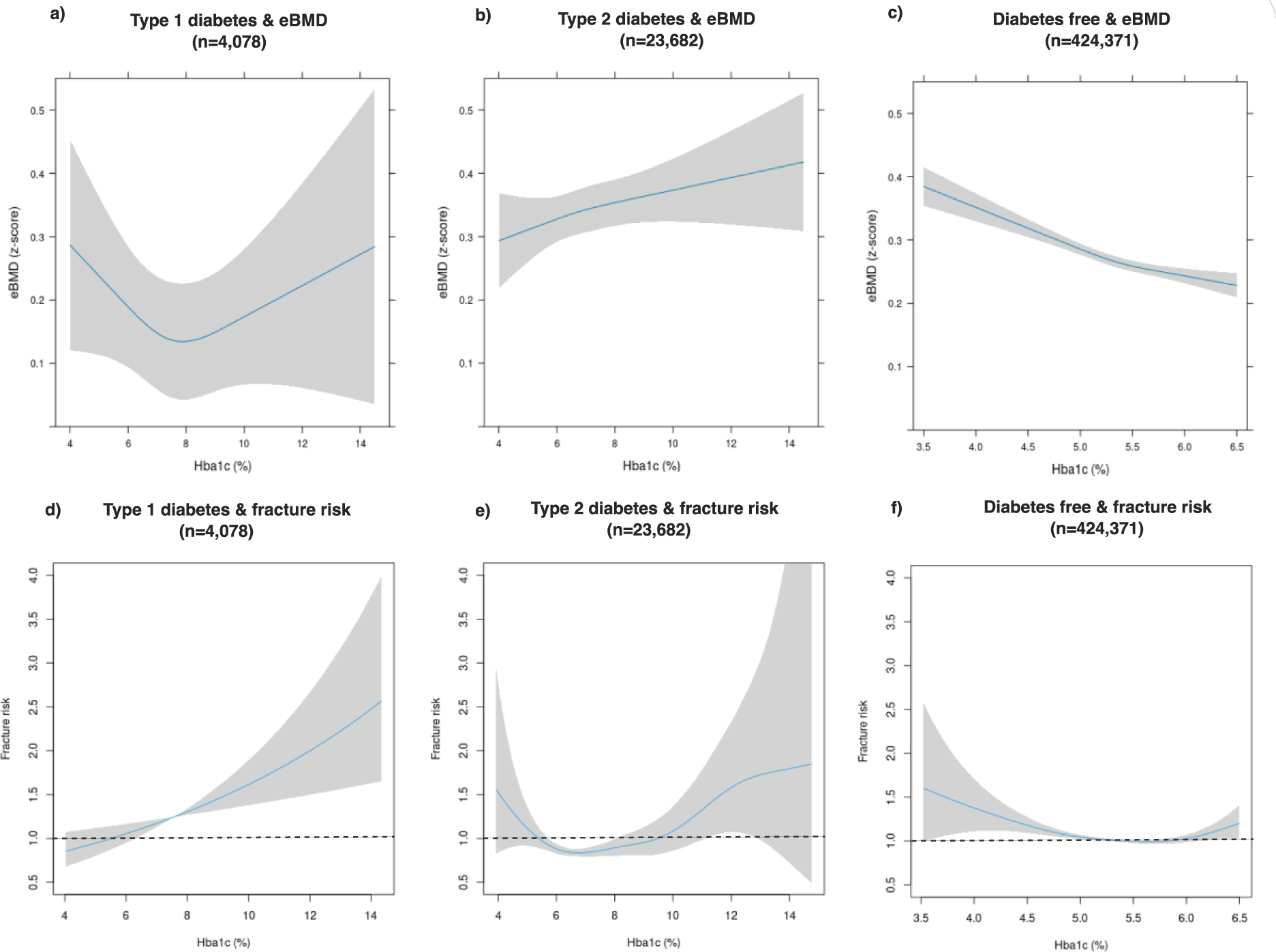
Associations between HbA1c and eBMD/fracture risk by diabetes status and type. Reference value was set to 5.4% (36mmol/mol) for all analysis groups. The shaded area represents the 95% confidence interval. For eBMD plots the covariates were set to the mean values. For example, for type 2 diabetes the covariates were: age=61, sex = male, weight=87.9 height=169, alcohol intake=1-2 per week. Creatinine=72.18 CRP=2.1, deprivation score=17.42, ethnicity = white European, duration of moderate activity=60, Non-smoker.

**Figure 2.**
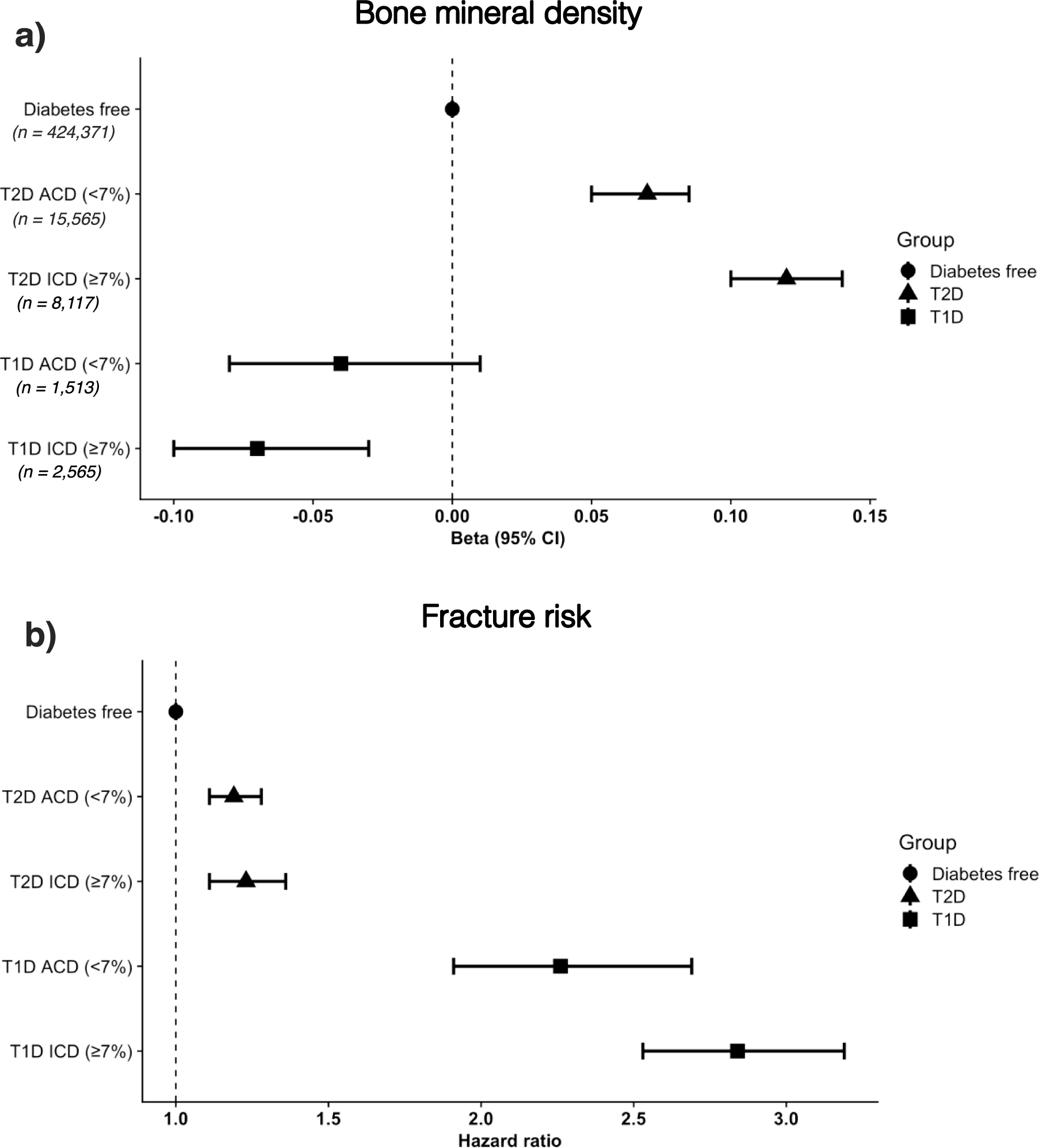
– a) Forest plot depicting associations between eBMD and glycemic control groups in comparison to controls. b) Forest plot depicting fracture risk ratios of glycemic control groups in comparison to controls. ICD = inadequately controlled diabetes, ACD = adequately controlled diabetes.

Individuals with ICD had increased eBMD in comparison to those with ACD (beta=0.04, 95%CI [0.01, 0.06]). In diabetes-free individuals, HbA1c levels were negatively associated to eBMD (beta=-0.05SD, 95%CI [-0.06, −0.04]). In addition, evidence for a non-linear association between HbA1c and eBMD was also observed (F-test ANOVA p-value = 0.02) (Figure1c).

### Glycemic control and fracture risk

A 1% (11mmol/mol) increase in HbA1c levels was associated with a 12% increase in fracture risk (HR: 1.12, 95%CI [1.05-1.19]) in individuals with type 1 diabetes (Figure 1d, supplementary table 3). Risk estimates were similar between men and women (supplementary table 4). In the type 1 diabetes sub-group analysis Cox regression Schoenfeld residual plots indicated a violation of the proportionality of hazards assumption for age as a covariate. However, upon conducting analyses stratified by age, with groups delineated as <38-50, <50-55, <55-60, <60-65, and <65-81 years, we found that the hazard ratios adhered to the proportionality assumption (supplementary Figure 1). Subsequently, associations between glycemic control groups and fracture risk were evaluated. As compared to individuals without diabetes, those with type 1 diabetes had increased fracture risk in both the ACD (HR 2.26, 95%CI [1.91, 2.69]) and ICD (HR 2.84, 95%CI [2.53, 3.19]) groups (Figure 2b). Further, individuals with ICD had a higher fracture risk than those with ACD (1.27, 95%CI [1.04, 1.57]).

Fracture risk in individuals with type 2 diabetes showed evidence for a U-shaped non-linear association with increased risk (F-test ANOVA p-value = 0.002) observed at both low and high levels of HbA1C (Figure 2e). A similar non-linear association was observed in diabetes-free individuals (F-test ANOVA p-value < 0.001), although attenuated within healthy HbA1c levels (Figure 2f). Duration of diabetes was found to be associated to fracture risk (supplementary Figure 2). However, when included in the type 2 diabetes Cox model no modification of the effect of HbA1c on fracture risk was observed. Fracture risk was increased in type 2 diabetes ACD (1.19, 95%CI [1.11, 1.28]) and ICD (1.23, 95%CI [1.11, 1.36]) groups, in comparison to diabetes-free individuals (Figure 2b). However, individuals with ICD did not have an increased fracture risk in comparison to those with ACD (0.97, 95%CI [0.91, 1.16]). Dichotomizing on a HbA1c level of 9% produced similar observations (supplementary Figure 3, supplementary table 3). We further split individuals with type 2 diabetes and ACD with a HbA1c level of lower than 5.4% and labelled this group as hypoglycemic (*n=*1,401). Individuals within the hypoglycemic group had the greatest fracture risk (1.49, 95%CI [1.20, 1.86]) (supplementary Figure 4). Additionally, individuals with pre-diabetes did not present higher fracture risk (HR: 0.99, 95%CI [0.96-1.03]; n=61,951) in comparison to controls.

### One-sample Mendelian randomization

One-sample Mendelian randomization analyses revealed no significant evidence of a linear or a non-linear association between genetically predicted HbA1c levels and fracture risk (power = 0.94) (supplementary Figure 5, supplementary table 6 & 7). With regards to eBMD, we observed significant evidence of a negative linear association between genetically predicted HbA1c levels and eBMD, in diabetes-free (beta=-0.09, 95%CI [-0.12, −0.05]) individuals and in the total population (beta=-0.08, 95%CI [−0.11, −0.05]) (supplementary Figure 6a, supplementary table 8). We observed no significant difference in effect estimates when related individuals were removed (supplementary table 8). Non-linear Mendelian randomization revealed strong non-linear relationship between HbA1c and eBMD in the overall population. The quadratic test yielded a p-value of 0.0002. This was supported by evidence from the fractional polynomial test (p-value = 0.009). Additional, evidence suggested that best-fitting fractional polynomial of degree 2 fitted the data better than the best-fitting fractional polynomial of degree 1 (p-value = 0.02). The best-fitting fractional polynomial of degree 2 had powers 1 and −1. The Cochran Q test was also significant (p-value = 0.001). Graphs and strata estimates indicated a negative slope from HbA1c levels between 4 - 6.5%, after which the slope becomes positive (supplementary Figure 6b, supplementary table 9, 10, 11). This relationship was not evident in analyses stratified by disease status (supplementary table 9). Yet, these analyses were underpowered (see supplementary tables 6 & 8 for power calculations).

## Discussion

We confirm an increased fracture risk in individuals with type 1 and type 2 diabetes from a relatively healthy young cohort [25]. Further, we observed a linear association between HbA1c and an increased fracture risk in individuals with type 1 diabetes and a non-linear association in individuals with type 2 diabetes. Individuals with type 1 diabetes and inadequately controlled diabetes (ICD, >7%/53mmol/mol) had greater fracture risk, than those with adequately controlled diabetes (ACD, < 7%/53mmol/mol). In type 2 diabetes, fracture risk did not differ between ACD and ICD groups even after increasing the HbA1c dichotomization cut-off to 9% (75mmol/mol). This lack of difference in fracture risk was observed despite higher BMD levels in individuals with ICD. However, individuals with type 2 diabetes and a HbA1c level below 5.4% (36mmol/mol) had the greatest fracture risk. In type 1 diabetes, we observed evidence that lowering HbA1c levels will reduce fracture risk. In type 2 diabetes, we observed evidence that both higher and lower HbA1c levels were associated with increased fracture risk. Our work brings to perspective the results of some previous studies [15, 26-29] indicating that individuals with type 2 diabetes and ACD present lower or similar fracture risk to that of diabetes-free individuals. Leveraging genetic information, we observed no association between genetically predicted higher HbA1c levels and fracture risk. However, we did observe evidence of a non-linear association between genetically-predicted HbA1c levels and eBMD in the total population. We observed a negative association between HbA1c and eBMD between 4.0 - 6.5% (20-48mmol/mol), after which the association to eBMD became positive. These results are concordant with our findings in the observational analysis and provide evidence of a causal non-linear relationship.

Previous studies have suggested that the increased fracture risk in type 2 diabetes is largely mediated by poor glycemic control [15, 26-29]. While individuals with type 2 diabetes and ICD showed a higher fracture risk compared to those without diabetes, the ACD group also had an increased fracture risk. No difference between ICD and ACD groups regarding fracture risk was observed, even after increasing the cut-off to 9% (75mmol/mol). Paradoxically, ICD was associated with higher eBMD, but this did not beget an expected decrease in fracture risk, a well-established phenomenon observed in individuals with type 2 diabetes [2]. Many of the studies reporting a significant relationship between poor glycemic control and fracture risk report varying cut-offs of HbA1c levels for defining adequate/inadequate glycemic control.

Clinicians often utilize cut-offs or threshold values to assist in the process of making decisions. However, their effectiveness is a subject of debate [30]. Cut-offs are problematic because they typically correspond to the specific population being studied and thus seldom produce consistent outcomes when applied to separate studies or datasets. Additionally, employing cut-offs to categorize a continuous predictor can potentially hinder accurate risk prediction [31]. Our results describe a U-shaped relationship between HbA1c and fracture risk in individuals with type 2 diabetes and diabetes-free individuals. Similarly, a large retrospective study of 652,901 elderly male veterans with type 2 diabetes observed that fracture risk was not increased in individuals with HbA1c > 8.5% (69mmol/mol), but was increased in individuals with HbA1c < 6.5% (48mmol/mol) [32]. Individuals with low levels of HbA1c can be considered as hypoglycemic [33]. Evidence suggests that hypoglycemia can lead to a loss of balance and an increase in falls, which can subsequently increase their fracture risk [34]. While this explanation seems plausible, it is important to state that the ADA suggests that HbA1c “does not provide a measure of glycemic variability or hypoglycemia” [31]. Alternatively, the observed association with increased fracture risk in individuals with very low HbA1c levels could be confounded by the use of glucose-lowering medication. However, this would not explain the increased risk in diabetes-free individuals. Individuals with anemia or other conditions shortening the lifespan of red blood cells (i.e., glucose-6-phosphate dehydrogenase deficiency, sickle-cell disease, etc.), may have underestimated HbA1c levels [35]. Considering the association between anemia and heightened risk of fractures [36], we incorporated measured hemoglobin concentrations into our statistical models to evaluate their impact. The inclusion of hemoglobin markedly enhanced the model’s explanatory power, as evidenced by a significant F-test ANOVA (p-value < 2×10^-16). Nevertheless, the estimated effects were not statistically different from those in the model that excluded hemoglobin concentrations (supplementary table 12). In type 1 diabetes, poor glycemic control was linearly associated with increased fracture risk. No significant differences in eBMD were observed across glycemic control groups. This could be a consequence of non-linear effects (for which we observed weak evidence) and/or low statistical power. In line with our findings, a nested case-control study in the UK reported no differences in fracture risk in relation to glycemic control among individuals with type 2 diabetes. However, similar to our study, they did observe a difference in fracture risk for those with type 1 diabetes [37]. Other studies also report increased fracture risk in type 1 diabetes individuals with poor glycemic control [14, 38]. Nevertheless, these studies reported different magnitude of effects likely due to differences in underlying cohort populations. Differences in the relationship between HbA1c and bone fragility across the different types of diabetes are expected. This can be a consequence of the distinct pathological mechanisms, age of disease onset and duration [39, 40]. Diabetes disease duration has been shown to affect both fracture risk and glycemic control in type 1 diabetes and type 2 diabetes [14]. However, when including duration as a covariate in our analysis, we observed no modification of the association between HbA1c and fracture risk.

Our study has some limitations proper to observational studies. Despite the large sample size, the low prevalence of incident fractures in this population means statistical power is still limited for the one-sample Mendelian randomization analyses of fracture risk. Power calculations for diabetes-specific stratified Mendelian randomization analyses suggest that a sample size of 218,351 individuals with diabetes is needed to achieve 80% power. We were unable to account fully for the consequences of diabetes disease duration and inadequate control in this study. Yet, controlling for diverse diabetes complications like diabetic retinopathy, kidney disease, and microvascular disease showed consistent effect estimates (supplementary table 12). Further, we employed HbA1c which was only measured at baseline and therefore was not reflective of long-term glycemic control. Additionally, the UK Biobank has been reported to suffer from ‘healthy volunteer bias’ [25]. As such, longitudinal studies with repeated measurements of HbA1c, a detailed assessment of diabetes duration, and evaluation of DXA-derived BMD change over time will provide a more comprehensive understanding of these associations. Additionally, prospective studies should consider examining other factors such as glycemic variability, hypoglycemic events, and the effects of glucose-lowering medications on fracture risk.

We observed distinct relationships between HbA1c and bone fragility in type 1 diabetes and type 2 diabetes and postulate that this is due to the underlying differences in disease pathology, including the anabolic effect of insulin on bone. We advise against the use of hard cut-offs for defining adequate/inadequate glycemic control as these can vary across the populations being studied. We obtained evidence that lower HbA1c levels will reduce fracture risk in patients with type 1 diabetes. In individuals with type 2 diabetes, lowering HbA1c levels can mitigate the risk of fractures up to a threshold, beyond which the risk may begin to rise once more. Our study contributes to the growing body of evidence on the relationship between glycemic control, fractures, and bone health, highlighting the need for individualized management strategies in individuals with diabetes to optimize skeletal outcomes and reduce fracture risk. Furthermore, longitudinal studies with repeated measurements are needed to identify the key determinants of fracture risk in type 2 diabetes.

## Data Availability

All data generated or analyzed during this study are included in this published article [and its supplementary information files].

https://magicinvestigators.org/downloads/

## List of abbreviations

(GWAS): Genome-wide association study
(LD): linkage disequilibrium
(type 2 diabetes): type 2 diabetes
(PRS): polygenic risk scores
(MR): Mendelian randomization
(SNP): single nucleotide polymorphism
(BMI): body mass index
(BMD): Bone mineral density
(DXA): Dual X-ray absorptiometry.

## Declarations

Not applicable.

## Ethics approval and consent to participate

All studies were conducted in full accordance with the ethical principles outlined in the Declaration of Helsinki, as revised in 2013, including adherence to protocols approved by their respective institutional ethics review committees and all participants provided written informed consent.

## Consent for publication

Not applicable.

## Availability of data and materials

HbA1C summary statistics can be obtained from MAGIC consortium website (https://magicinvestigators.org/downloads/). All data generated or analyzed during this study are included in this published article [and its supplementary information files].

## Authors’ contributions

S.Ghatan, L.Oei, F.Rivadeneira, designed the study. S.Ghatan performed the analysis. S.Ghatan, L.Oei, F.Rivadeneira, C.Medina-Gomez, F.Koromani, and K.trajanoska drafted the manuscript. All authors contributed to the interpretation of data and critical revision of the manuscript. All authors read and approved the final version of the manuscript.

## Acknowledgments

John Kemp for providing assistance in coding and quality control of the estimated bone mineral density data. This research has been conducted using the UK Biobank Resource (accession ID: 67864). We express our gratitude to the study participants, whose participation made this work possible, and to the numerous colleagues who contributed to the collection, phenotypic characterization of clinical samples, as well as genotyping and analysis of GWAS data. S.Ghatan is the guarantor of this work and, as such, had full access to all the data in the study and takes responsibility for the integrity of the data and the accuracy of the data analysis.

## Originality and Prior Publication

Parts of the results from this manuscript have been presented previously at the European Calcified Tissue Society (ECTS) 2023 congress in Liverpool, England on 16^th^ of April.

