## Supplementary figures for "Evaluating the relationship between glycemic control and bone fragility within the UK biobank: Observational and one-sample Mendelian randomization analyses"

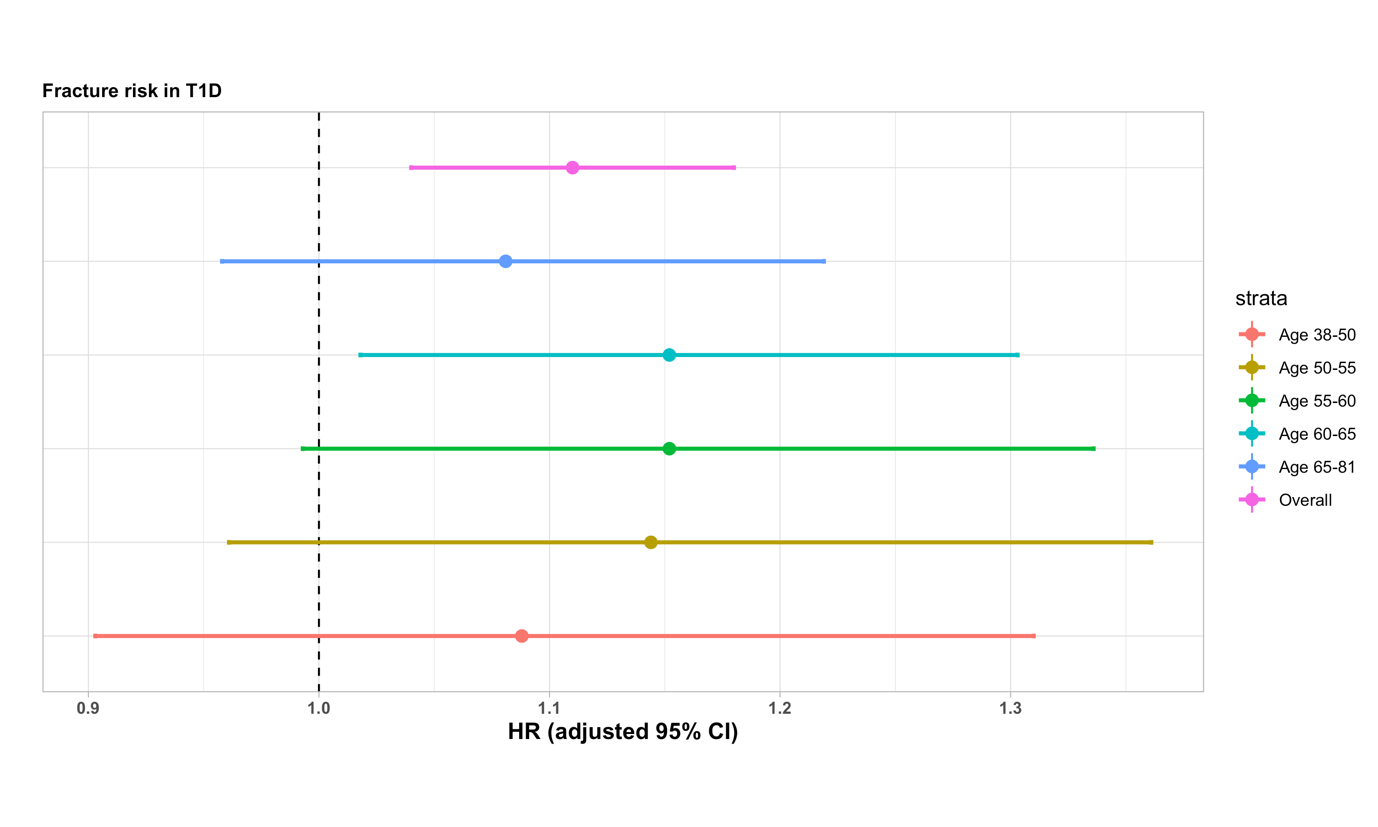
Supplementary figure 1 – Age stratified association between Hba1c and fracture risk in T1D

Supplementary figure 2 – Association between diabetes duration and fracture risk.

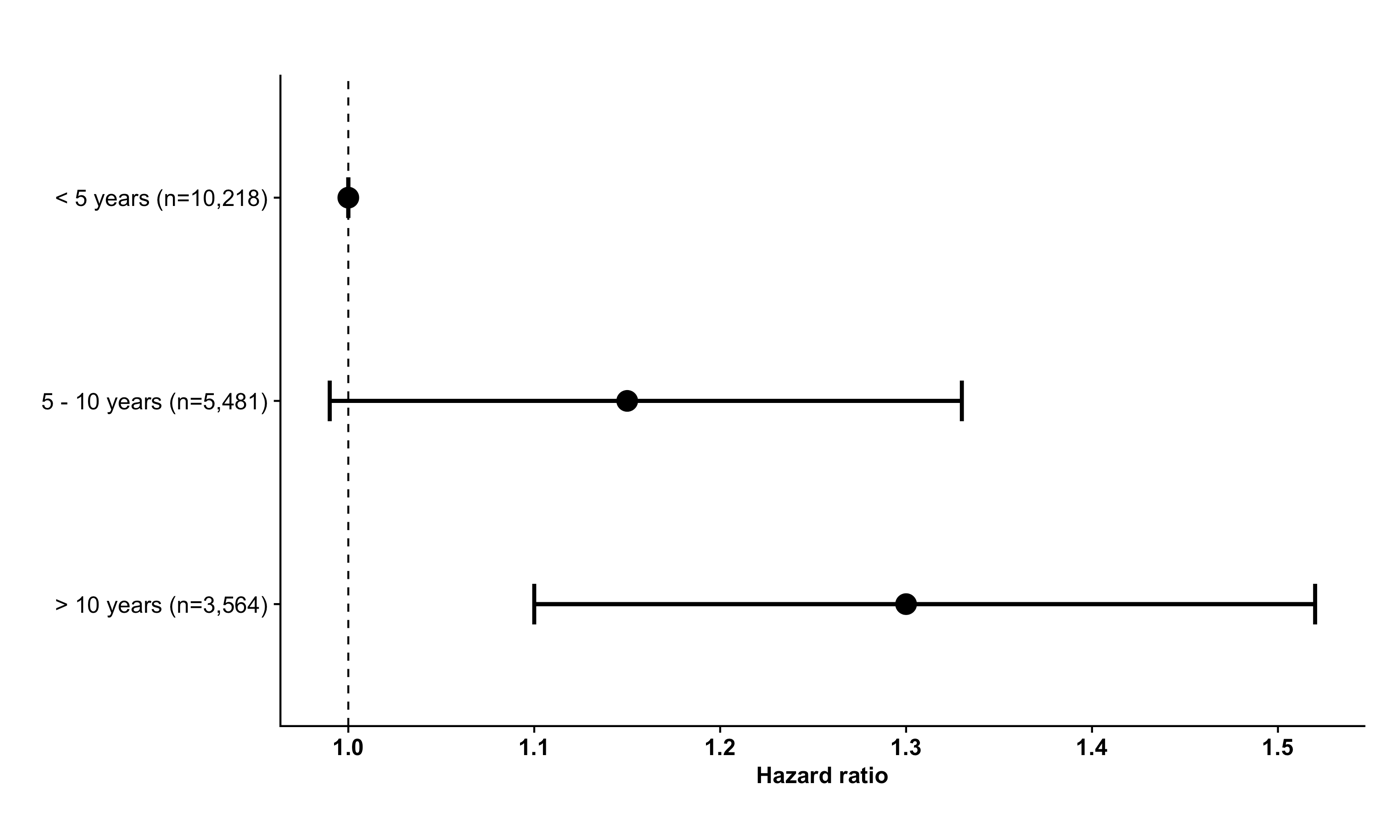

Supplementary figure 3 – Forest plot depicting fracture risk hazard ratios of glycemic control groups in comparison to controls. ICD = inadequately controlled diabetes, ACD = adequately controlled diabetes.

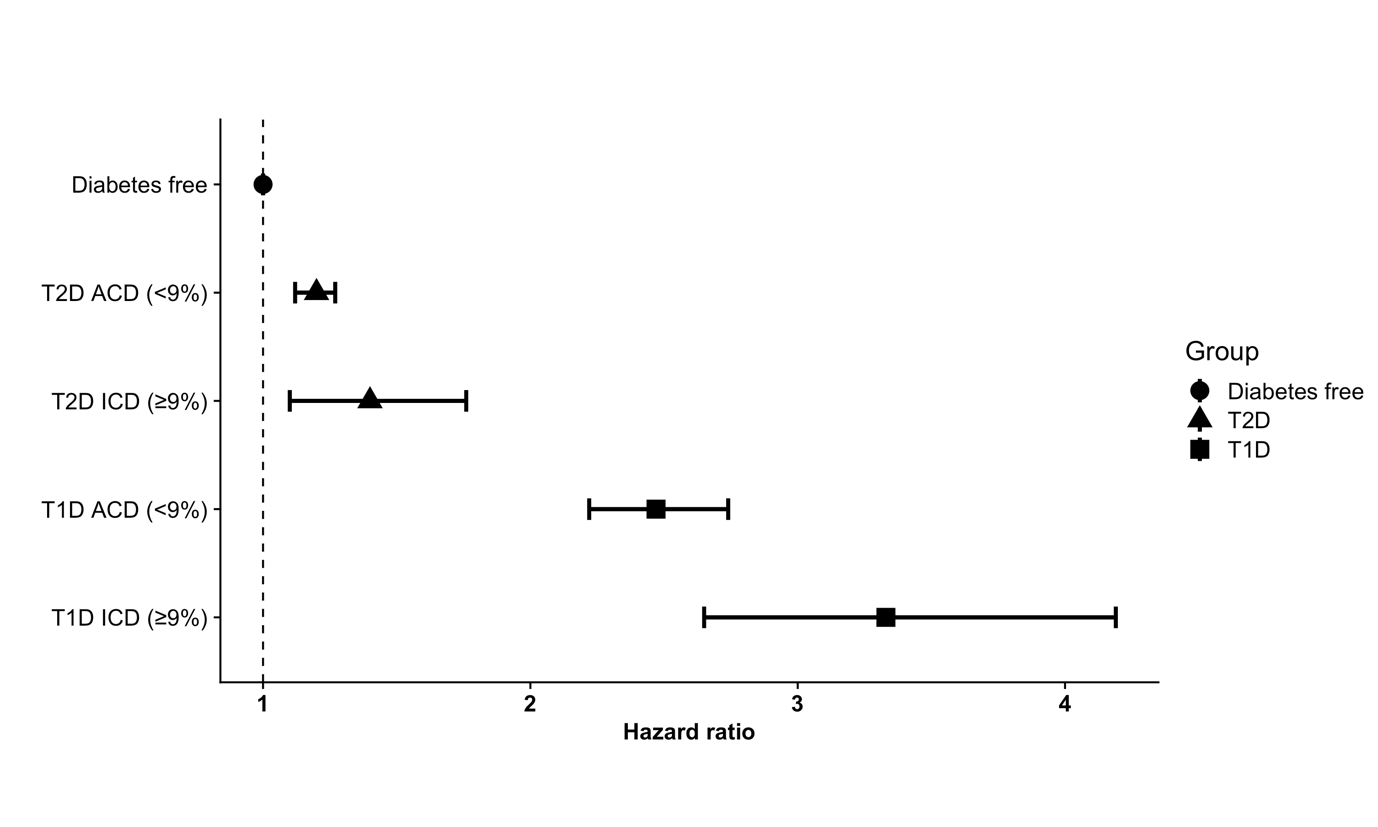

Supplementary figure 4 - Forest plot depicting fracture risk hazard ratios of T2D glycemic control groups in comparison to controls. ICD = inadequately controlled diabetes, ACD = adequately controlled diabetes.

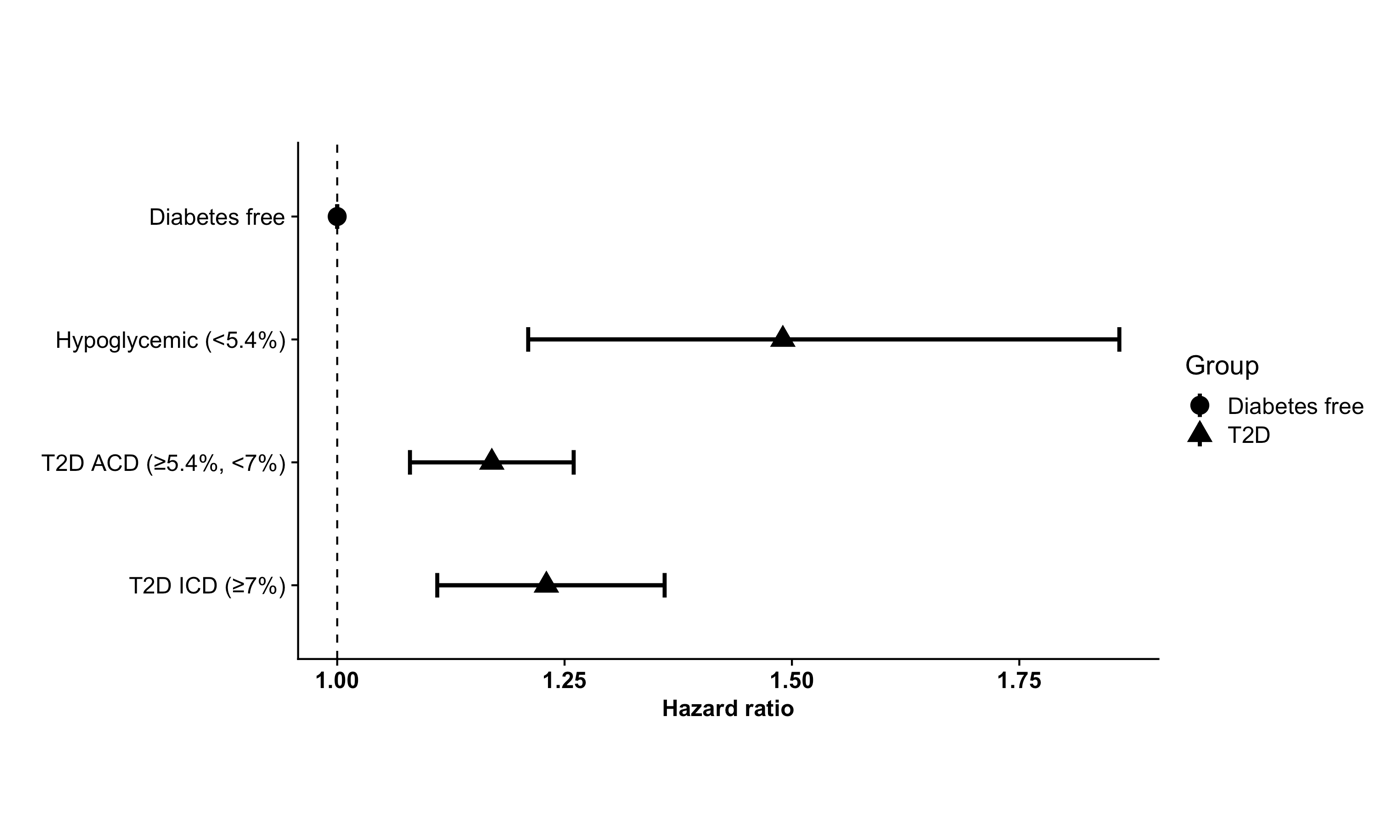

Supplementary figure 5 – Forest plot depicting Mendelian randomisation results between HbA1C and fracture risk.

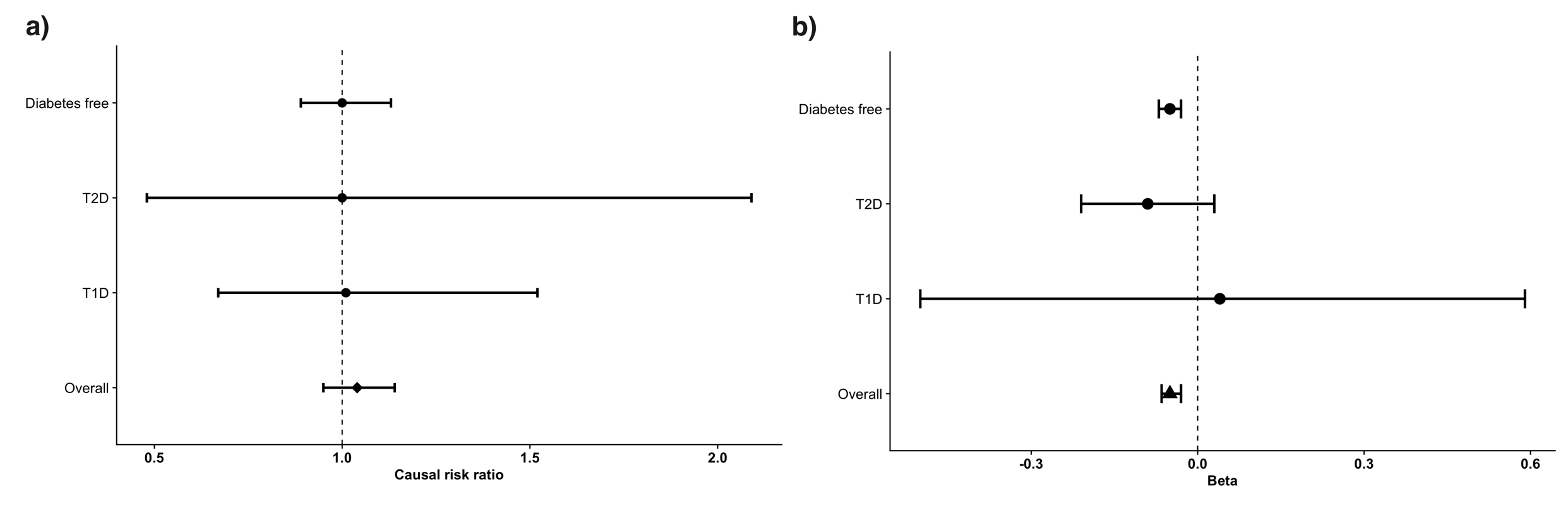

Supplementary figure 6 – a) Linear Mendelian randomization between HbA1c and eBMD (b) Non-linear Mendelian randomization using fractional polynomial method. The red dot represents the reference level 5.4% (36mmol/mol). Grey lines 95% confidence intervals. Number of strata 100.

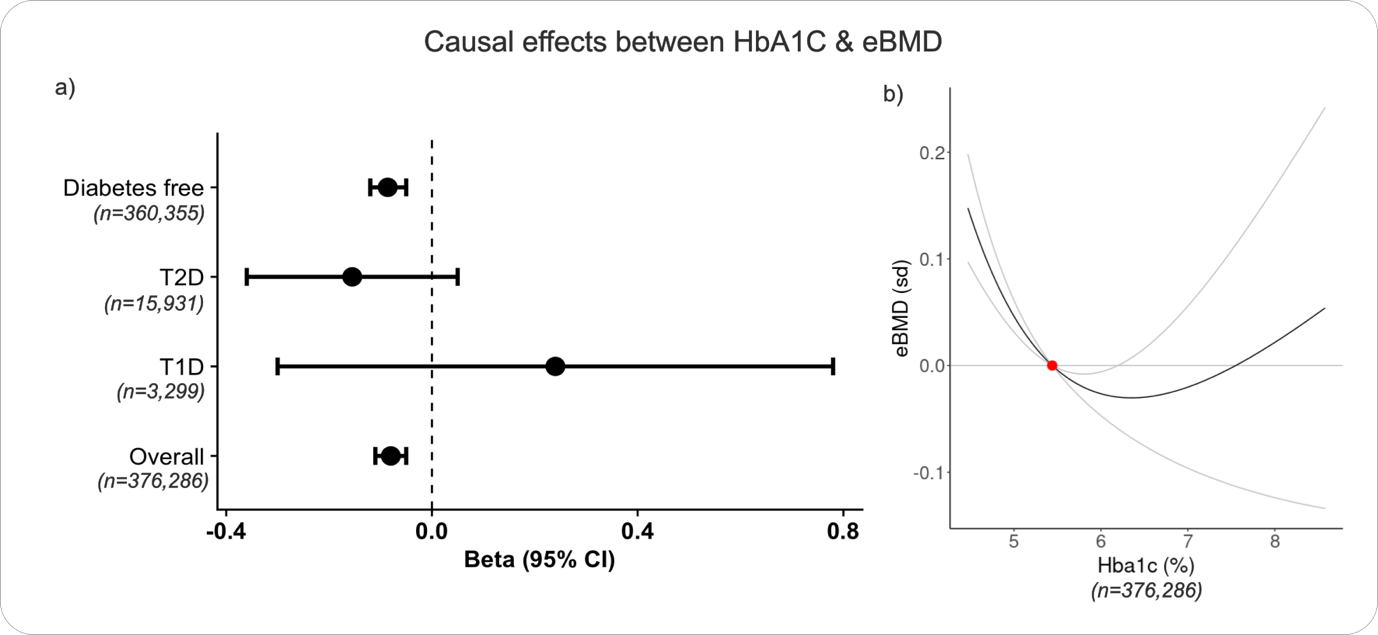
